# Neurological Imaging Order Selection Using Natural Language Processing and a Support Vector Classifier

**DOI:** 10.1101/2023.06.24.23291863

**Authors:** Videet Mehta, Rohan Dharia, Nilesh Desai

## Abstract

**Purpose:** To develop an algorithm for automated medical imaging order selection based on provider-input signs and symptoms using natural language processing and machine learning. The aim is to reduce the frequency of inappropriate physician imaging orders, which currently accounts for 25.7% of cases, and thereby mitigate potential patient health concerns.

**Materials and Methods:** The study was conducted retrospectively with a four-step analysis process. The data used for training in the study consisted of anonymized imaging records and associated provider-input symptoms for CT and MRI orders in 40,667 patients from a tertiary children’s hospital. First, the data were normalized using keyword filtering and lemmatization. Second, an entity-embedding ML model converted the symptoms to high-dimensional numerical vectors suitable for model comprehension, which we used to balance the dataset through k-nearest-neighbor-based synthetic sampling. Third, a Support Vector Classifier (ML model) was trained and hyperparameter-tuned using the embedded symptoms to predict modality (CT/MRI), contrast (with/without), and anatomical region (head, neck, etc.) for the imaging orders. Finally, a web application was developed to package the model, which analyzes user-input symptoms and outputs the predicted order.

**Results:** The model was found to have a final overall accuracy of 93.2% on a 4,704-case test set (*p <* 0.001). The AUCs for the eight classes ranged from 96% to 100%, and the average F1-score was 0.92.

**Conclusion:** This algorithm looks to act as a clinical decision support tool to help augment the present physician imaging order selection accuracy and improve patient health.

## 1 Introduction

Magnetic resonance imaging (MRI) and computed tomography (CT) are routinely used in the diagnosis of various diseases, playing a crucial role in disease identification for millions of people worldwide [1]. These imaging modalities allow healthcare professionals to visualize and identify pathologies, and physicians can then carefully plan treatments for the abnormalities accurately.

### 1.1 Purpose

Although their general purpose is the same, MRI and CT scans are necessary for different diagnosis requirements. MRI utilizes non-ionizing nuclear magnetic resonance to generate three-dimensional images of bone and soft tissue structures [2]. In contrast, CT scans employ multiple X-ray projections to produce high-resolution cross-sectional images of bone and soft tissues [3]. These modalities offer unique advantages and limitations for specific research applications involving different pathologies and anatomical regions. Accurate differentiation between the two seemingly different imaging modalities and their respective suitable applications is crucial. The complexity of an image order, including factors like location and the use of contrast, contributes to the intricacy of the process, thereby increasing the likelihood of ordering errors by physicians.

### 1.2 Concerns

Imaging appropriateness has been of important concern in the effort to improve medical imaging efficiency and cost reduction. A previous study indicated that 26% of medical imaging orders were deemed inappropriate, signifying the selection of an incorrect scan rather than the appropriate one [4]. This high proportion has four main consequences for patients. (i) Re-imaging leads to excessive radiation [5] ii) MRI scans especially require the youngest of pediatric patients or those with developmental delay to be sedated. Repeated or prolonged sedation has short-term risks such as cardiopulmonary risks and potential neuro-cognitive long-term risks [6] (iii) Diagnosis may be delayed, incomplete, or mischaracterized (iv) Repeat imaging has cost implications for patients, healthcare systems, and insurance providers [7].

Clinical decision support (CDS) systems have been suggested to improve the efficiency of imaging ordering and reduce re-imaging rates significantly [8]; however, there is a lack of comprehensive research available to assert their effectiveness. Furthermore, existing CDS acts solely to verify physician decisions rather than provide an independent second opinion, and with it, appropriateness concerns persist [4, 9]. As an alternative approach to this problem, we conducted a study to develop an AI algorithm with NLP that can independently determine the most appropriate imaging order for a patient based on their symptoms in a clinical setting.

## 2 Methods

The retrospective study aimed to develop a classification model that improves imaging order selection accuracy by providing the most appropriate imaging order for any given entry of free-text data. This model serves as an add-on to existing imaging order processes.

### 2.1 Data

Data were obtained from a Hospital’s Department of Radiology following IRB approval. Inclusion criteria were patients with CT/MRI examinations performed at the campus from 01/01/2010-10/11/2022. No exclusion criteria were used.

All imaging orders were verified for accuracy and anonymized per HIPAA guidelines through manual data review by employees prior to the data’s use in the study. The class labels (imaging orders) within the dataset were thus considered the ground truth for training and testing.

The dataset contained 40,667 entries of patient data (maximum available given inclusion criteria), including the imaging order, as seen in Table 1. Exploratory data analysis was first conducted to identify key features for implementation in the algorithm.

**Table 1:**
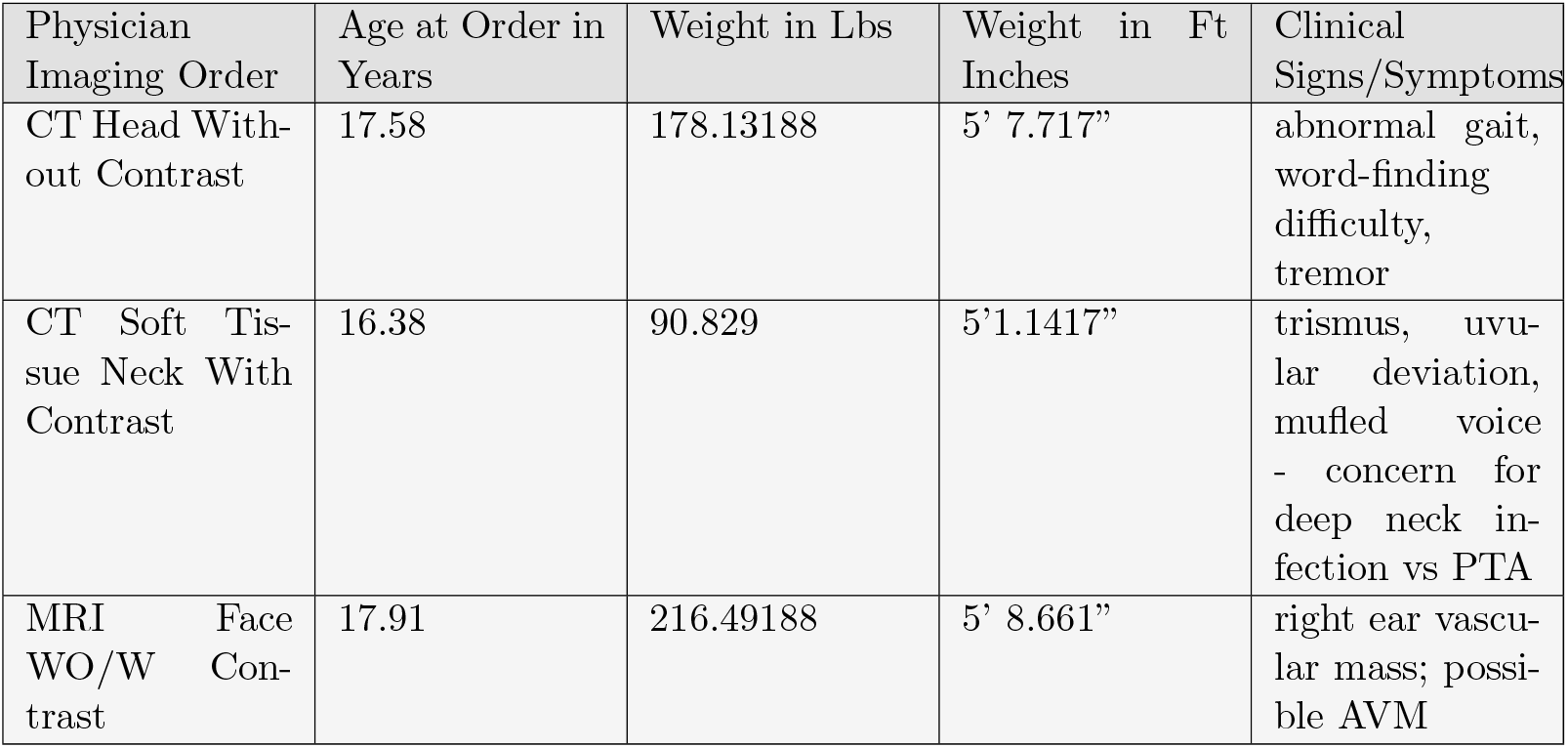
Sample Data.

| Physician Imaging Order | Age at Order in Years | Weight in Lbs | Weight in Ft Inches | Clinical Signs/Symptoms |
| --- | --- | --- | --- | --- |
| CT Head Without Contrast | 17.58 | 178.13188 | 5’ 7.717” | abnormal gait, word-finding difficulty, tremor |
| CT Soft Tissue Neck With Contrast | 16.38 | 90.829 | 5’1.1417” | trismus, uvular deviation, muffled voice - concern for deep neck infection vs PTA |
| MRI Face WO/W Contrast | 17.91 | 216.49188 | 5’ 8.661” | right ear vascular mass; possible AVM |

Among the data columns were various unidentifiable patient attributes such as age, weight, and height, which were removed due to low predictive power. Missing and irregular data was handled by removing any rows with empty or exclusively non-alphanumeric symptom text.

Following data analysis, the predictor variable was defined as the physician’s manually written symptom text, due to the absence of significant alternative predictive features. The outcome variable was the imaging order required for the patient.

### 2.2 Preprocessing

A five-step processing algorithm addressed data irregularities.

The initial step was to handle outliers, which were defined as cases with imaging orders with *<* 900 total instances in the dataset [10]. The final distribution of imaging order frequencies following the removal of these orders can be seen in Fig. 3b.

**Figure 1.**
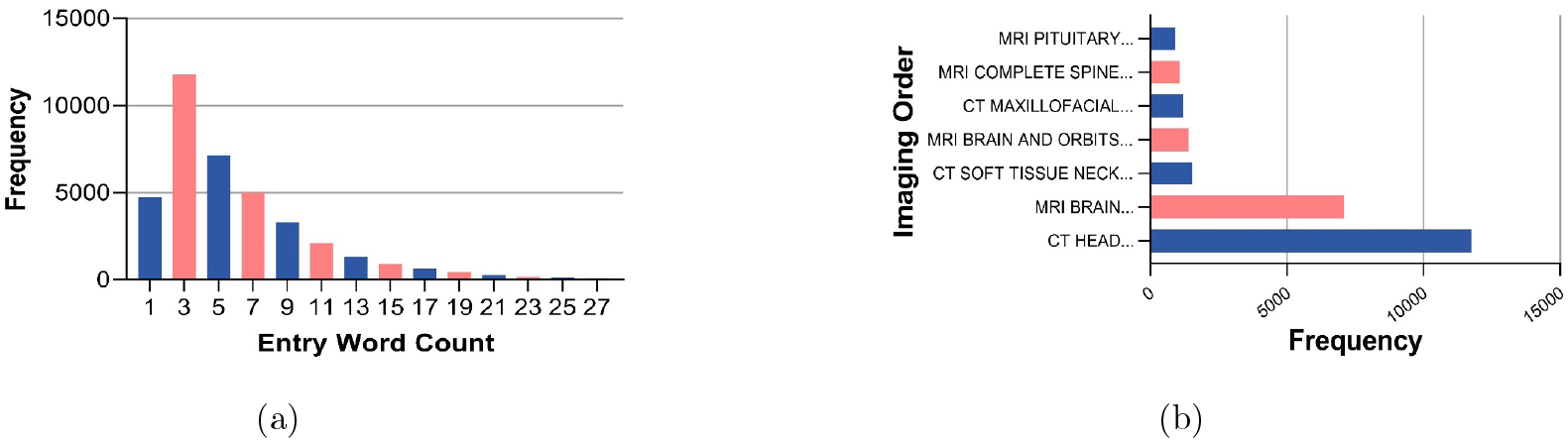
Exploratory data analysis of the dataset of (**??**) frequency of entry word count and (**??**) frequency of each imaging order following the removal of outliers.

**Figure 2.**
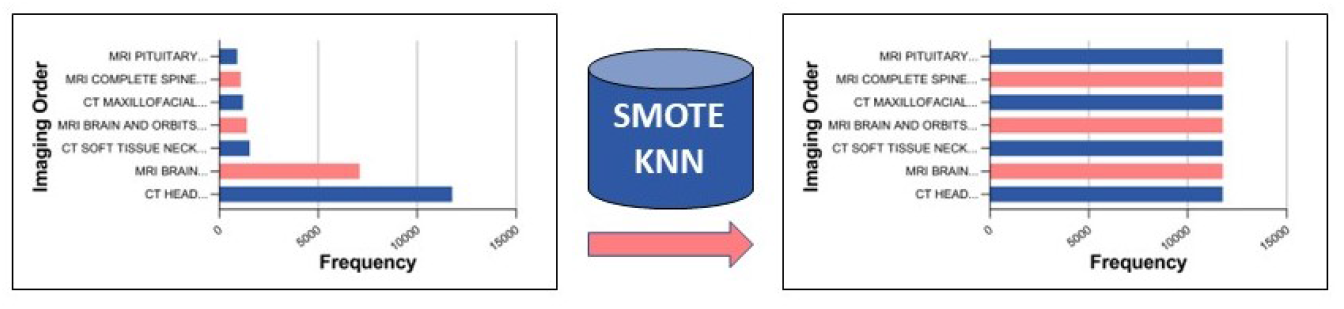
Diagram shows the SMOTE k-nearest-neighbor-based oversampling technique used to counter the class-imbalance. Left graph shows an uneven class distribution prior to SMOTE and right graph shows an even distribution of 11,760 data entries per class after. SMOTE = Synthetic minority oversampling technique

**Figure 3.**
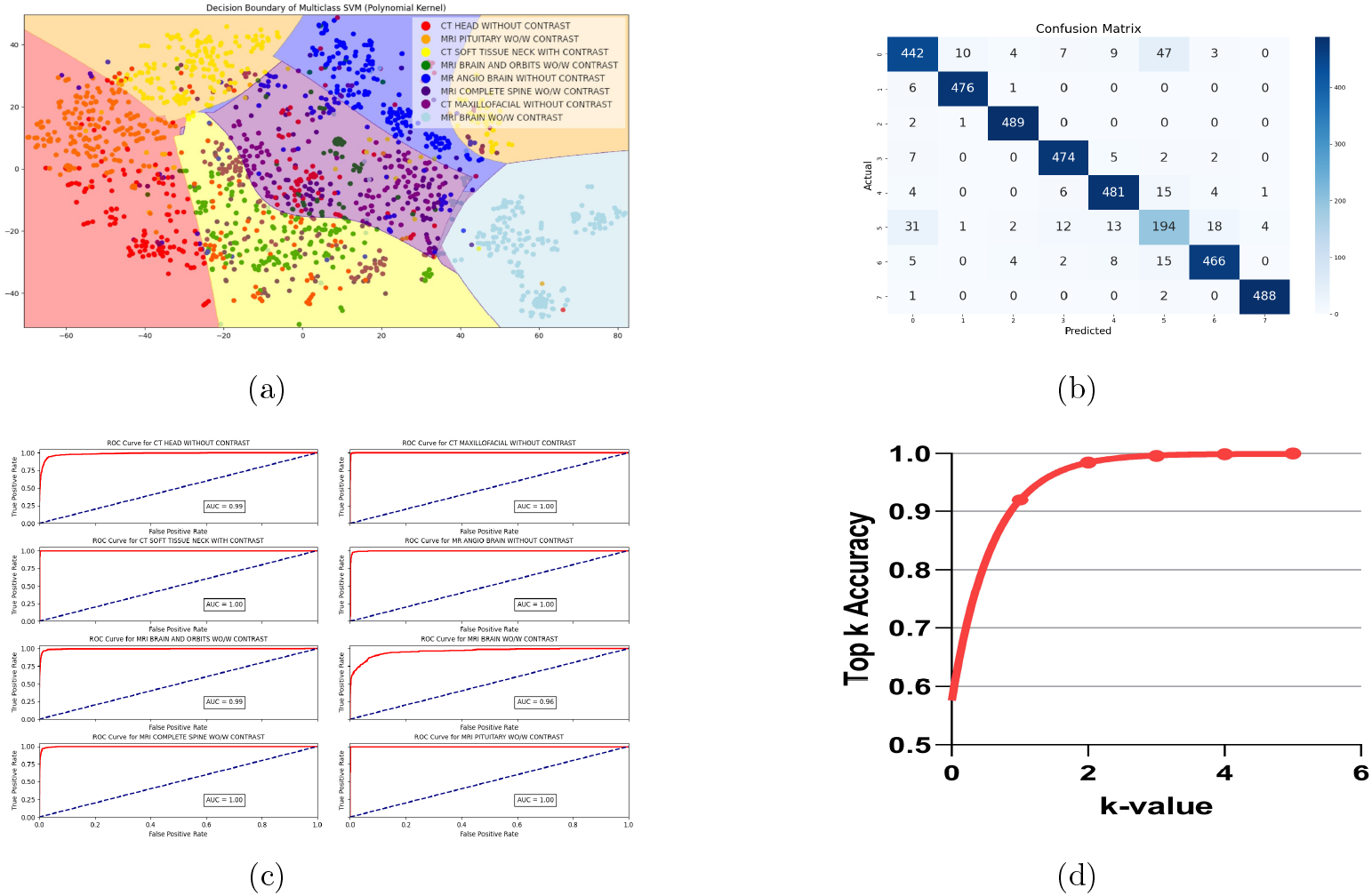
Graphs show **(a)** t-distributed stochastic neighbor embedding (t-SNE) visualization of decision boundaries drawn by Support Vector Classifier (SVC), with each color representing a distinct class and each plot representing a datapoint, **(b)** confusion matrix of SVC model on test set (n=4704), with darker colors representing the increased overlap between model predictions and class labels, **(c)** receiver operating characteristic (ROC) curves on individual class predictions with associated areas under the curve (AUC), demon-strating tradeoff between sensitivity and specificity for each imaging order, and **(d)** Top-K curve displaying accuracy of the correct class being in the top K predictions for each value K between 1 and 5 (reaching 100%).

To condense the symptom text, the Stopword corpus from Python’s Natural Language Toolkit (NLTK) removed any insignificant words [11]. Word lemmatization then simplified each word to its most fundamental grammatical root in order to reduce noise for the model [12].

To transform the text into numerical representations, we leveraged OpenAI’s 1.2 billion-parameter pre-trained Ada embedding model, which uses word embedding techniques to convert each symptom text row into dense, 1536-dimensional numerical vectors conveying both the definition and context of the input [13, 14].

To combat a high class imbalance (Fig. 3b), a synthetic minority oversampling technique (SMOTE) k-nearest-neighbor algorithm increased the number of samples of the minority classes and grew the overall sample size from 40,667 to 94,080 rows (Fig. 2) [15, 16].

### 2.3 Data Partitions

Of the 94,080-case sample size, 95% of the data was used for training, and 5% (4,704 cases) became an unbiased testing group. This low testing split was chosen due to the high training data requirements of NLP tasks [10]. All data were partitioned randomly at the patient level with no systematic differences between groups.

### 2.4 Model

The models tested were XGBoost, an algorithm that optimizes a loss function using gradient boosting and decision parallel tree construction; Random Forest, an ensemble learning method that constructs multiple decision trees and aggregates their outputs; and Support Vector Classifier (SVC), which identifies a hyperplane to optimally separate data points of different classes in high-dimensional feature space [17]. The models were implemented from Python’s scikit-learn v1.1.2 library. A nested One-vs-All classifier generated 8 separate models for each algorithm to evaluate class-specific metrics.

### 2.5 Training

Each model was trained on the training dataset using default hyperparameters apart from a random state of 42. Solely raw accuracy based on model performance on the testing dataset was measured for preliminary evaluation. Accuracy, in terms of true positive (TP), true negative (TN), and sample size (N) is defined as:

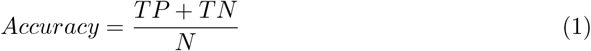

The model with the highest initial accuracy on the testing set underwent hyperparameter tuning using GridSearchCV to maximize final accuracy.

### 2.6 Evaluation

The final model was evaluated with the testing set, which originated from the same data source as in training but was necessary due to data limitations. For this study, false negative and false positive cases carried equal penalty, so F1 scores, the harmonic mean of precision and recall, best indicated the model’s ability to predict each imaging order individually [18, 19].

To further evaluate the model, a top-K accuracy chart, receiver-operating characteristic (ROC) curves, and confusion matrices were also created. A one-way Student’s t-test then validated the model by comparing it to the physician ordering method using a P value of 0.001 for reference.

## 3 Results

The SVC exhibited the highest accuracy of 91.5% on the test partition, outperforming the other models (XGBoost = 84.6%, Random Forest = 82.3%). Table 2 reports each model’s respective accuracies. The GridSearchCV then increased the SVC accuracy by 1.7% to a final accuracy of 93.2% through hyperparameter tuning. The final hyperparameters were the polynomial kernel, C = 9, and gamma = 2.38. This accuracy statistic is superior to the 74.3% accuracy rate in physicians in selecting the appropriate medical imaging order (*P <* 0.001). Further, the predict probabilities parameter reports the probability of each class being the correct imaging order, allowing for physician independence to decide the correct imaging order in the case of disagreements or uncertainty by the model. Fig. 3d reports a 100% accuracy in the correct imaging order being listed in the model’s top three predictions.

**Table 2:** Classification Report.

| Imaging Order | Count | Precision | Recall | F1 |
| --- | --- | --- | --- | --- |
| CT HEAD WITHOUT CONTRAST | 585 | 0.85 | 0.83 | 0.84 |
| CT MAXILLOFACIAL WITHOUT CONTRAST | 588 | 0.95 | 0.99 | 0.97 |
| CT SOFT TISSUE NECK WITH CONTRAST | 577 | 0.97 | 0.99 | 0.98 |
| MR ANGIO BRAIN WITHOUT CONTRAST | 585 | 0.94 | 0.98 | 0.96 |
| MRI BRAIN AND ORBITS WO/W CONTRAST | 603 | 0.94 | 0.94 | 0.94 |
| MRI BRAIN WO/W CONTRAST | 557 | 0.84 | 0.72 | 0.77 |
| MRI COMPLETE SPINE WO/W CONTRAST | 617 | 0.91 | 0.95 | 0.93 |
| MRI PITUITARY WO/W CONTRAST | 590 | 0.97 | 0.99 | 0.98 |
| Sum/Average | 4704 | 0.92 | 0.92 | 0.92 |

A tSNE-dimensionality reduction technique to compress the 1536-numerical vectors along with visualizing the decision boundaries of the SVC is shown in Fig. 3a. Although extremely simplified, we see several distinct regions of clustered points, indicating a high classification accuracy for those specific imaging orders.

The ROC Curves shown in Fig. 3c demonstrate a strong tradeoff between sensitivity and specificity in the steep rises shown for each imaging order class. The AUC’s ranged from 0.96 to 1.00, indicating the model reaches high true positive rates without sacrificing precision and with relatively low false positive rates. However, MRI Brain WO/W CONTRAST (AUC: 0.96) had the lowest AUC scores.

As seen from the confusion matrix in Fig. 3b, the model’s inaccuracies were most frequently found in confusion between the MRI BRAIN WO/W CONTRAST and CT HEAD WITHOUT CONTRAST classes.

## 4 Discussion

This tool utilizes an AI-based NLP approach to improve physician accuracy in medical imaging order selection, increasing it from approximately 74% to around 93%. The algorithm was developed by leveraging patient symptom data to accurately predict the most appropriate imaging orders. Existing CDS tools, such as CareSelect imaging, utilize clinical guidelines to assess whether certain diagnostic tests are necessary for a given patient. However, these tools fail to independently predict the most appropriate test and provide a second opinion for the physician; instead, they simply verify existing physician decisions [9]. This pipeline aims to provide an additional, evidence-based position to foment the most positive patient outcomes.

To facilitate widespread implementation, the developed pipeline has been incorporated into a user-friendly web application. This application accepts free-text symptom input and generates the predicted imaging order along with a confidence score. Further advancement will need to involve integrating the algorithm directly into physician software.

The widespread implementation of this algorithm holds potential for several positive outcomes as a result of decreased re-imaging due to more accurate imaging order selection [8]. First, excessive radiation brought by several imaging methods, namely CT scans, would be mitigated and radiation exposure would be reduced to a minimal level.[20, 21] The algorithm would, furthermore, eliminate repeated sedation, especially in pediatric patients, reducing the potential for complications [22]. Reducing re-imaging would also expedite patient treatment timelines, as the process of imaging, including carrying out the imaging, reading results, and conveying the outcome to the patient, is time-consuming, and for patients with unknown critical conditions, repeating these steps due to physician imaging order error can be life-threatening [23]. Implementing this algorithm for precise imaging ordering offers a significant advantage of cost reduction for both hospitals and patients [23]. By avoiding excessive imaging orders, hospital systems can save millions of dollars, leading to improved patient care [24].

Despite the numerous benefits of this tool, there exist several avenues for further development that would optimize its effectiveness. Enhancement of this algorithm’s accuracy metrics will most importantly require the incorporation of more training data. The present data includes just one feature (clinical symptoms) and originates from a single hospital institution. Incorporating vital signs, laboratory orders, family history, and clinical guidelines from multiple institutions in the model could certainly see an additional 4-5% accuracy increase.

The present dataset also is limited to neurological imaging orders; however, due to the generalizability of the algorithm, this can be easily diversified [25]. Because this model does not need to be adapted to specific features of the dataset, any additional, similarly-structured data (free-text input + class-based output) could be used in training to also produce high accuracy rates. In this way, the scope of the model could quickly be grown to fit any medical specialty, and the application’s online nature will allow for real-time remote updates to the platform. A final area of future development is the integration of clinical guidelines into the algorithm’s processing. Similar CDS tools have seen success with the consideration of, for example, the ACR Appropriateness Criteria, in the evaluation of imaging orders. Similar attention in future CDS imaging order models would not only improve accuracy but also account for continuously evolving literature and medical knowledge.

In conclusion, this tool will allow physicians to significantly improve the accuracy of their medical imaging orders with an easily accessible approach using solely a text-based patient evaluation. As a result, users would see a decrease in costs, radiation exposure, sedation, and delays due to the decreased frequency of repeated imaging. This solution will look to remedy the high levels of inappropriate orders currently seen in the medical field, and future development will further contribute to that goal.

## Data Availability

The data provided in the manuscript is a private dataset from Texas Childrens Hospital not available for request.

